# Global and local ancestry modulate *APOE* association with Alzheimer’s neuropathology and cognitive outcomes in an admixed sample

**DOI:** 10.1101/2022.02.02.22270331

**Authors:** Michel Satya Naslavsky, Claudia K. Suemoto, Luciano Abreu Brito, Marília Oliveira Scliar, Renata Eloah Ferretti-Rebustini, Roberta Diehl Rodriguez, Renata E. P. Leite, Nathalia Matta Araujo, Victor Borda, Eduardo Tarazona-Santos, Wilson Jacob-Filho, Carlos Pasqualucci, Ricardo Nitrini, Kristine Yaffe, Mayana Zatz, Lea T. Grinberg

**Affiliations:** Departamento de Genética e Biologia Evolutiva, Instituto de Biociências, University of São Paulo, São Paulo, Brazil; Human Genome and Stem Cell Research Center, University of São Paulo; Hospital Israelita Albert Einstein, São Paulo, Brazil; Division of Geriatrics, University of São Paulo Medical School, Sao Paulo, Brazil; Escola de Enfermagem, Programa de Pós-Graduação em Enfermagem na Saúde do Adulto, University of São Paulo, São Paulo SP, Brazil; Department of Neurology, University of São Paulo Medical School, Sao Paulo, Brazil; Department of Pathology, LIM-22, University of São Paulo Medical School, Sao Paulo, Brazil; Departamento de Genética, Ecologia e Evolução, Instituto de Ciências Biológicas, Federal University of Minas Gerais, Belo Horizonte, Brazil; University of Maryland School of Medicine; Department of Psychiatry, Neurology, and Epidemiology and Biostatistics, University of California, San Francisco, San Francisco, USA; VA Medical Center, San Francisco, San Francisco, USA; Memory and Aging Center, Department of Neurology, and Pathology, University of California, San Francisco, San Francisco, USA; Global brain Health Institute, University of California, San Francisco, San Francisco, USA

## Abstract

Dementia is more prevalent in Blacks than in Whites, likely due to a combination of environmental and biological factors. Paradoxically, clinical studies suggest an attenuation of *APOE* ε4 risk of dementia in African ancestry (AFR), but lack of neuropathological data preclude the interpretation of the biological factors underlying these findings, including the association between *APOE* ε4 risk and Alzheimer’s disease (AD) pathology, the most frequent cause of dementia. We investigated the interaction between African ancestry, AD-related neuropathology, *APOE genotype*, and functional cognition in a postmortem sample of 400 individuals with a range of AD pathology severity and lack of comorbid neuropathology from a cohort of community-dwelling, admixed Brazilians. Increasing proportions of African ancestry (AFR) correlated with a lower burden of neuritic plaques (NP). However, for individuals with high levels of NP and neurofibrillary tangles (NFT), AFR proportion was associated with worse Clinical Dementia Rating sum of boxes (CDR-SOB). Among *APOE* ε4 carriers, the association between AFR proportion and CDR-SOB disappeared. *APOE* local ancestry inference of a subset of 309 individuals revealed that, in *APOE* ε4 noncarriers, non-European *APOE* background associated with lower NP burden, but with worst cognitive outcomes compared to European *APOE* when adjusting by the similar NP burden. Finally, *APOE* ε4 was associated with worse AD neuropathological burden only in a European *APOE* background. *APOE* genotype and its association with AD neuropathology and clinical pattern are highly influenced by ancestry, with AFR associated with lower NP burden and attenuated *APOE4* risk compared to European ancestry.

## Introduction

Recent data from the USA suggest a prevalence of Alzheimer disease (**AD**)-type dementia (**AD dementia**) of 19% in non-Hispanic Blacks compared to 10% in non-Hispanic Whites, with Hispanics having a prevalence in between these estimates ^1, 2^. A study from a large UK cohort also suggest that older Blacks have a higher prevalence of dementia than Whites ^3^ Paradoxically, several studies suggest that apolipoprotein E ε4 allele (***APOE*4**), considered the strongest genetic risk factor for late-onset AD, has a weaker effect in Blacks than in Whites ^4, 5^ or that the effect of *APOE*4 on AD risk is attenuated in African ancestry (**AFR**), especially when the *APOE* gene is on an African local ancestry ^6-8^. Indeed, *APOE*4 allele frequency varies across populations, consistently more common in AFR than European ancestry (**EUR**) (12-21% vs. 6-14%, respectively) ^9, 10^. Factors related to genetic ancestry architecture, specific traits, or disease-related genetic variants enriched in a particular population group due to founder effects may provide biological explanations for these differences ^11, 12^. However, pinpointing the biological impact of ancestry-related genetic architecture in AD risk has been challenging. Self-reporting race is a poor predictor of ancestry-related biological effects because social determinants of health influence it, i.e., environmental factors present where people live and work that impact disease risk ^13-18^. Also, self-reported race is a qualitative metric, whereas genetic-based ancestry is quantitative, thus providing a better metric of admixture. More recent work in AD using genomic-based analysis of ancestry instead of self-reported race has advanced the field. However, the impact of most of these work is still limited by the lack of neuropathological confirmation^6, 19^, since non-White individuals are still underrepresented in autopsy studies. Neuropathological examination is the gold standard to diagnose and score age-related neuropathological findings. Most aging individuals show multiple comorbid neuropathological changes that impact clinical outcomes. Unfortunately, antemortem diagnosis remains unavailable to most of these conditions, including TDP-43 proteinopathy, synucleinopathies, and aging-related tau astrogliopathies ^20, 21^. This limitation hampers the ability to investigate ancestry-related biological differences, as opposed to differences due to social determinants of health, in AD risk or to determine whether the *APOE*4 risk in AD is really attenuated in AFR or the lower risk of dementia is related to increased amounts of non-AD neuropathology in individuals with AFR. The latter is possible. In a previous study investigating ancestry-driven differences in risk of developing neuropathological changes commonly associated with dementia, we used a panel of ancestry-informative markers (**AIMs**) in a cohort of 202 autopsy cases of individuals dwelling in São Paulo, a highly admixed 12 million inhabitants city in Brazil. We found that subjects with higher proportions of AFR ancestry showed a lower prevalence of neuritic plaques (OR=0.43, 95% CI=0.21–0.89, p=0.02) than individuals with EUR ancestry, when adjusted for sociodemographic variables and *APOE* genotype. Conversely, other neuropathological alterations, such as AD-tau burden, Lewy body disease, or microvascular brain changes, showed similar prevalence among different ancestries^22^.

To interrogate the correlation between proportion of AFR, AD neuropathology, clinical decline, and *APOE*4 risk, we have expanded our original clinicopathological cohort sixfold (1,333 individuals over 50 years of age) to obtain the necessary power of analysis to investigate only cases showing evidence of AD-related neuropathological changes (N=400). We excluded individuals with other non-AD neuropathological changes to avoid confounders in the analysis. We interrogated whether the percentage of AFR correlates with functional cognitive scores in participants stratified by *APOE4 status* over equivalent AD-neuropathology burden. Finally, as disadvantaged social determinants of health are enriched in particular ancestry groups, biological and environmental contributions must be considered when interpreting results associated with global ancestry. To isolate a possible ancestry-related biological effect, we investigated the interaction between AD neuropathological burden, cognition, and AFR, stratifying individuals by their local ancestry within the *APOE* locus.

## Methods

### Participants

A full-body autopsy is mandatory in Sao Paulo city when an individual dies from undiagnosed non-traumatic death. This study, which the local ethics committee approved, included samples and data from the Biobank for Aging Studies (BAS) from the University of Sao Paulo Medical School (Brazil) that were collected between 2004 and 2017 23, 24. The BAS includes individuals who were 18 years and older at the time of death and with informants who had at least weekly contact with the deceased in the six months prior to death. Exclusion criteria for participating in the BAS cohort included inconsistent clinical data or brain tissue unsuitable for neuropathological analyses (cerebrospinal fluid pH <6.5 or acute brain lesions that required examination by the pathologist in charge for the cause of death certification). A detailed explanation of the BAS procedures can be found elsewhere ^24^.

Trained gerontologists perform clinical and functional assessments. After the informed consent had been signed, the most knowledgeable informant was interviewed to obtain the deceased’s clinical history using a semi-structured interview, which has shown good evidence of validity for detecting cognitive impairment by informants in postmortem settings ^25 26^. Cognitive impairment was assessed using the Clinical Dementia Rating Scale (**CDR**) – informant section ^27^.

For this study, we only included participants lacking neuropathological changes (normal controls) or those with evidence of AD-related neuropathological changes (plaques and/or neurofibrillary tangles), but only if they lacked non-AD neuropathological lesions. We excluded cases with incomplete clinical, neuropathological, and genetic data.

### Genetic analyses and variables

DNA samples were obtained from blood or brain tissue and genotyped using Illumina OmniExpress 700k microarray or Illumina BeadXpress custom genotyping panel, as detailed in **Supplementary Methods**. *APOE* common alleles were preferably genotyped directly using allele-specific amplification ^28^ or after imputation of rs429358 to compose haplotypes (detailed in **Supplementary Methods**). Individuals were classified as either *APOE*4 carriers (at least one ε4 allele) or noncarriers, hereby defining the *APOE*4+ or *APOE4-* status, respectively. Global tri-hybrid continental ancestry was inferred using 47 ancestry informative markers (AIMs) with Structure 2.3.4 ^29^ using K=3. The panel is a subset from a previously described panel ^22^, which combined HapMap, Human Genome Diversity Project (**HGDP**), and New York Cancer Project (**NYCP**) as parental populations that was used to calibrate our inference (see **Supplementary Methods**). Global ancestry was analyzed in two ways: dichotomously, using a 2% cutoff for AFR (first quartile of the AFR distribution)^22^; and as a continuous variable, in increments of 10% AFR proportions indicating the presence of the African component in admixed individuals.

In the 309 individuals with available genotype microarray data, local ancestry inference (LAI) using large reference panels that included Brazilians and Brazilian parental populations were used to determine local ancestry of each *APOE* allele, as African, European, and Native American ^30^ (detailed in **Supplementary Methods**). For analysis involving local *APOE* ancestry, the 309 individuals were first classified into *APOE*4+ (at least one ε4 allele, n=74, **group A**) and *APOE*4- (only ε3 and/or ε2 alleles, n=235, **group B**). Given the predominance of European alleles, the *APOE*4*+* group was further divided into four groups: European *APOE*4+ (**group A1**, n=31, including homozygotes), mixed ancestry with one European *APOE*4+ and one non-European *APOE*4*-* allele (“Mixed *APOE*4+ 1” **group A2**, n=3), mixed ancestry with one non-European *APOE*4+ and any European *APOE*4- allele (“Mixed *APOE*4+ 2” **group A3**, n=23) and non-European *APOE*4+ (**group A4**, n=17, including homozygotes). The *APOE*4- was further divided into three groups: European *APOE*4- (**group B1**, n=138), mixed ancestry *APOE*4- (**group B2**, n=73), and non-European *APOE*4- (**group B3**, n=24). **Figure 1** and **Supplementary Table 1** provide all counts of individuals per *APOE* genotypes and local ancestries.

**Table 1.** Characteristics of the sample (n=400)

|  | < 2%AFR<br>n=100 | ≥ 2%AFR<br>n=300 | p |
| --- | --- | --- | --- |
| **Age (years), mean (SD)\*** | 73.4 (11.3) | 71.6 (12.4) | 0.20 |
| **Female, %<sup>†</sup>** | 47.0 | 51.3 | 0.45 |
| **Education (years), mean (SD)\*** | 5.9 (4.0) | 4.3 (3.7) | <0.001 |
| **Reported race/ethnicity, %<sup>‡</sup>** |  |  | <0.001 |
| *White* | 97.0 | 60.0 |  |
| *Black* | 0 | 16.7 |  |
| *Brown* | 3.0 | 22.3 |  |
| *Asian* | 0 | 1.0 |  |
| **CDR ≥ 0.5, %<sup>†</sup>** | 18.0 | 15.0 | 0.48 |
| **CDR sum of boxes, mean (SD)\*** | 1.7 (4.8) | 2.1 (5.2) | 0.76 |
| **At least one APOE ε4, %<sup>†</sup>** | 25.0 | 27.7 | 0.60 |
| **Braak & Braak score, %<sup>†</sup>** |  |  | 0.56 |
| *0-II* | 68.0 | 65.0 |  |
| *III-IV* | 21.0 | 26. |  |
| *V-VI* | 11.0 | 9.0 |  |
| **CERAD score, %<sup>†</sup>** |  |  | 0.09 |
| *None or Sparse* | 72.0 | 77.0 |  |
| *Moderate* | 10.0 | 13.0 |  |
| *Frequent* | 18.0 | 10.0 |  |

AFR: African global ancestry; SD: standard deviation; CERAD: Consortium to Establish a Registry for Alzheimer's disease
\*unpaired t-test; <sup>‡</sup>Fisher exact test; <sup>†</sup>chi-square test

**Figure 1.**
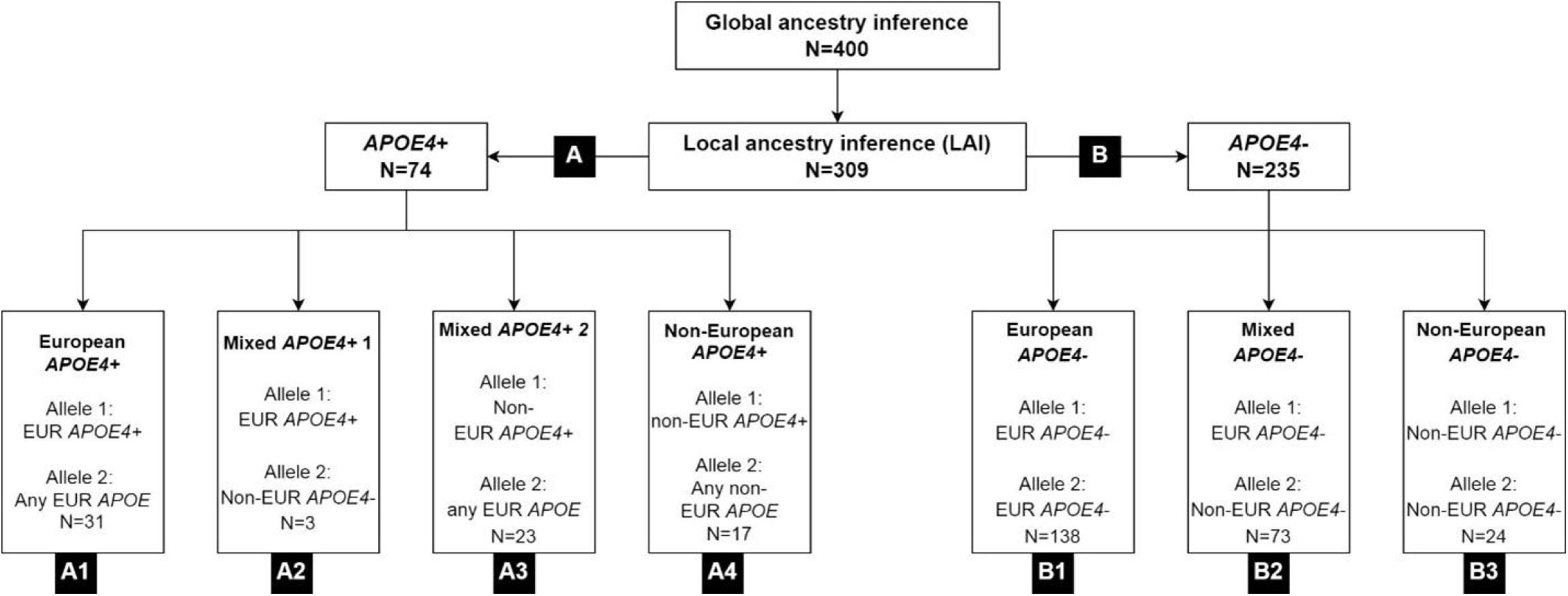
Groups of individuals defined by *APOE* genotype and local ancestry inference (n=309). While groups A1, A4, B1, and B3 contain individuals with a single local ancestry at APOE locus, groups A2, A3, and B2 contain individuals with dual ancestry at the APOE locus (mixed). Group A APOE4 carriers, group B are composed of noncarriers.

### Neuropathological assessment

The BAS uses a 14-region immunohistochemistry panel to detect neurodegeneration using universally accepted criteria to stage and diagnose cases^24^. Neurofibrillary tangle (**NFT**) pathology was scored by Braak stage, encoded into four groups following the conventional categorization ^31^: Braak stages 0, I/II, III/IV, and V/VI. β-amyloid plaque pathology was scored using Consortium to Establish a Registry for Alzheimer’s Disease (**CERAD**) for the density of neuritic plaques, scored as none, sparse, moderate, or frequent.

### Postmortem interview

The clinical interview included information on age, sex, and reported race by the informant, the latest confirmed by the interviewer on a government-issued document with photo identification. The informant also reported on the deceased’s years of formal education.

Cognitive function was evaluated using the CDR scale, which is a five-point scale used to classify dementia severity in six domains: memory, orientation, judgment and problem solving, community affairs, home, and hobbies, and personal care ^27^ The CDR was designed to be applied to both the patient and the informant, but due to study design, only the informant part was used. As the study outcome, we used the CDR sum of boxes (CDR-SOB, range: 0 to 18).

### Statistical analysis

We used descriptive statistics to examine the characteristics of the sample by AFR status, categorized into two groups defined by a cutoff of 2% of AFR. This cutoff was defined based on the AFR proportion present in 90% of Caucasian samples from the Human Genome Diversity and HapMap Projects in 2012 ^32,22^. We used independent groups Student’s t-test for continuous variables and chi-squared or Fisher’s exact tests to describe categorical variables. Subsequent analysis classified individuals based on the proportion of AFR ancestry (measured as a continuous 10% increasing scale). We determined the association of AD-neuropathology burden (Braak stages and CERAD scores) with AFR using linear regression models adjusted for age, sex, and education. We further adjusted this model for *APOE*4 status (*APOE*4+ vs. *APOE*4-). Variance inflation factor (**VIF**) values did not indicate multicollinearity between Braak stages and CERAD scores in our sample.

We investigated the association between AFR and cognition, independent of AD-neuropathology burden, using the CDR-SOB as a continuous outcome and linear regression models adjusted for Braak stages and CERAD scores. These models were adjusted for age, sex, education, and *APOE*4 status.

Next, we examined whether AFR was an effect modifier in the relationship between AD-neuropathology burden and CDR-SOB scores by creating interaction terms of AFR with Braak stages and CERAD scores. Since we hypothesized that *APOE*4 could modify the previous interactions, we tested the hypothesis of a triple interaction between AFR, AD-neuropathology, and *APOE*4. Further, we stratified the previous interaction analyses by *APOE*4 status.

Finally, we investigated the association between CDR-SOB and the local ancestry of *APOE* using linear regression models adjusted for age, sex, and education. As indicated in tables, AD-neuropathology scores were used as outcomes or CDR-SOB covariates, and models were adjusted for AFR. Groups based on local *APOE* ancestry (**Figure 1**) were used both in *APOE4-* individuals to test local ancestry effects (group B1 vs. B2+B3; B1 vs. B3, see **Results**) and between *APOE4+* vs. *APOE4-* individuals of the same local ancestry (groups A1 vs. B1 and A4 vs. B3, respectively, see **Results**) to test allele-specific effects for each context. All analyses used Stata 15 (StataCorp, College Station, TX). The alpha level was set at 0.05, and all tests were two-tailed.

## Results

### Demographics

The analysis included 400 participants with a mean age of 72.0 ± 12.1 years old. 50% were women, the mean education attainment was 4.7 ± 3.9 years, and 27% of the participants had at least one *APOE*4 allele. Participants were reported as White (69%), Brown (18%), and Black (12%). Genomic-based ancestry analysis revealed that 75% of participants had ≥ 2% of AFR ancestry, a group that contains a significant proportion of White (65%), most Brown, and all Black individuals. Of note, few individuals had >80% AFR even among those reported as Black, highlighting the admixed composition of this cohort. In univariate analyses, the ≥ 2% AFR group had similar age and sex distribution but a lower educational level than the < 2% AFR group (**Table 1**). The ≥ 2% AFR group had a greater proportion of individuals with a lower plaque burden. However, we detected no differences in CDR score close to death, *APOE*4 status, or neurofibrillary tangles burden between the two groups. (**Table 1**). The average AFR proportions within individuals with declared White, Brown, and Black ethnicity groups were 11%, 36%, and 61%, respectively, demonstrating a broad range of admixture (**Supplementary Figure 1 and Supplementary Table 1**).

### Analysis using dichotomous ancestry grouping

We replicated the analysis of our 2013 paper ^22^ in this larger cohort with 400 subjects with pure AD neuropathology or lacking significant neuropathological changes and obtained similar results (**Supplementary Table 2**). Participants with ≥2% AFR had lower CERAD scores than those with < 2%AFR (β= -0.210, 95% CI=-0.418; -0.001, p=0.04). As before, Braak stage showed no association with an ancestry group.

**Table 2.**
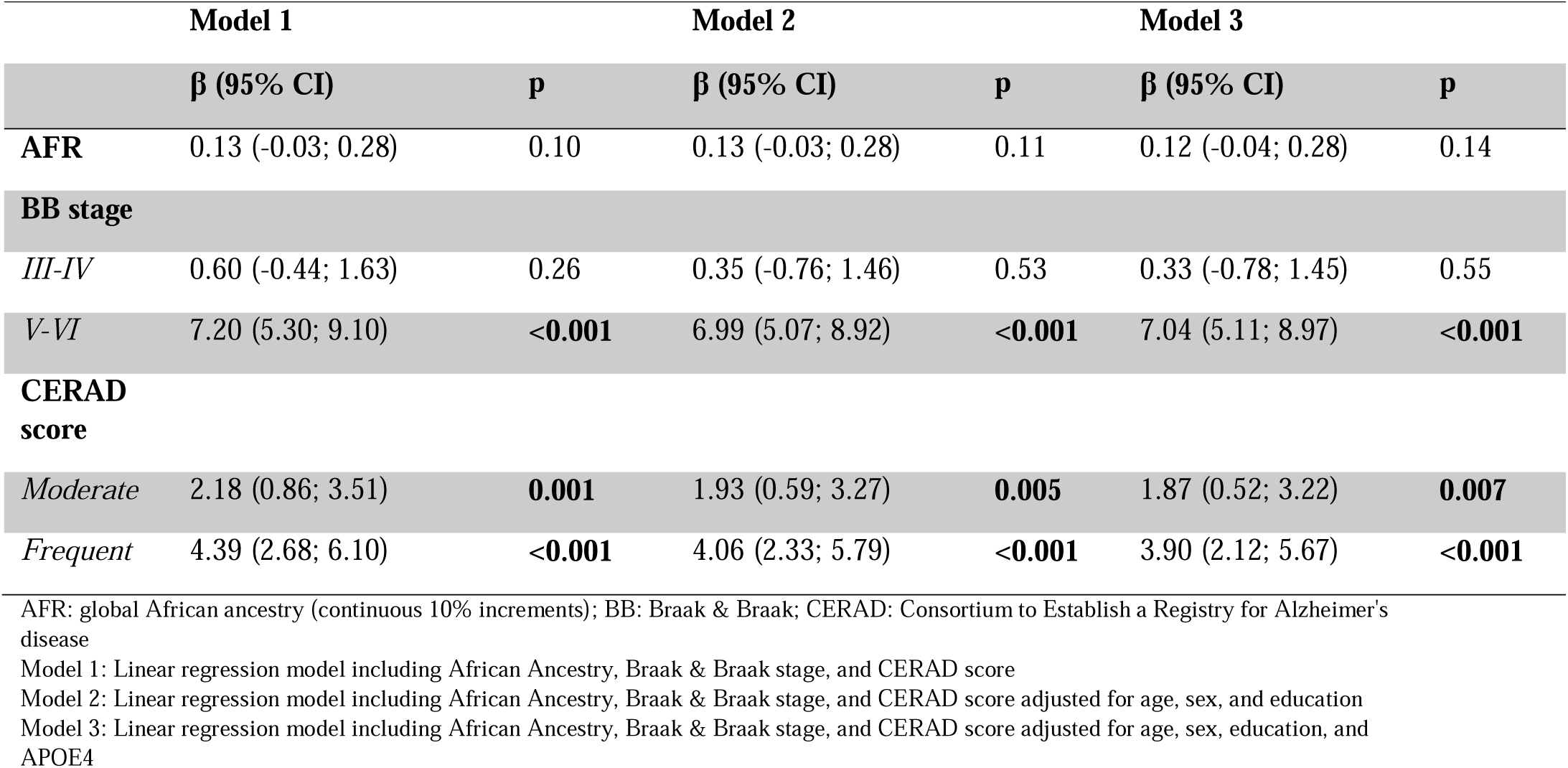
Association of Clinical Dementia Rating sum of boxes (CDR-SOB) with quantitative proportions of African ancestry and AD neuropathological burden (n=400)

|  | Model 1 |  | Model 2 |  | Model 3 |  |
| --- | --- | --- | --- | --- | --- | --- |
| | $\beta$ (95% CI) | p | $\beta$ (95% CI) | p | $\beta$ (95% CI) | p |
| **AFR** | 0.13 (-0.03; 0.28) | 0.10 | 0.13 (-0.03; 0.28) | 0.11 | 0.12 (-0.04; 0.28) | 0.14 |
| **BB stage** |  |  |  |  |  |  |
| *III-IV* | 0.60 (-0.44; 1.63) | 0.26 | 0.35 (-0.76; 1.46) | 0.53 | 0.33 (-0.78; 1.45) | 0.55 |
| *V-VI* | 7.20 (5.30; 9.10) | **<0.001** | 6.99 (5.07; 8.92) | **<0.001** | 7.04 (5.11; 8.97) | **<0.001** |
| **CERAD score** |  |  |  |  |  |  |
| *Moderate* | 2.18 (0.86; 3.51) | **0.001** | 1.93 (0.59; 3.27) | **0.005** | 1.87 (0.52; 3.22) | **0.007** |
| *Frequent* | 4.39 (2.68; 6.10) | **<0.001** | 4.06 (2.33; 5.79) | **<0.001** | 3.90 (2.12; 5.67) | **<0.001** |

AFR: global African ancestry (continuous 10% increments); BB: Braak & Braak; CERAD: Consortium to Establish a Registry for Alzheimer's disease Model 1: Linear regression model including African Ancestry, Braak & Braak stage, and CERAD score Model 2: Linear regression model including African Ancestry, Braak & Braak stage, and CERAD score adjusted for age, sex, and education Model 3: Linear regression model including African Ancestry, Braak & Braak stage, and CERAD score adjusted for age, sex, education, and APOE4

### Analysis using quantitative African ancestry proportions

Using this larger cohort, we obtained increased analysis power to interrogate whether the increasing proportion of AFR modifies the association between AD-neuropathology and cognitive abilities. We found no correlation between AFR and functional cognitive abilities (CDR-SOB score as the outcome) (**Table 2**).

We found significant interactions between AFR and AD-neuropathology (p-value for AFR interaction with CERAD=0.007; and Braak=0.002) (**Figure 2A and 2B**). This suggests that, among individuals with severe AD neuropathology, the higher the AFR proportion, the worse the CDR-SOB scores are, even after adjustment for age, sex, education, and *APOE* status.

**Figure 2.**
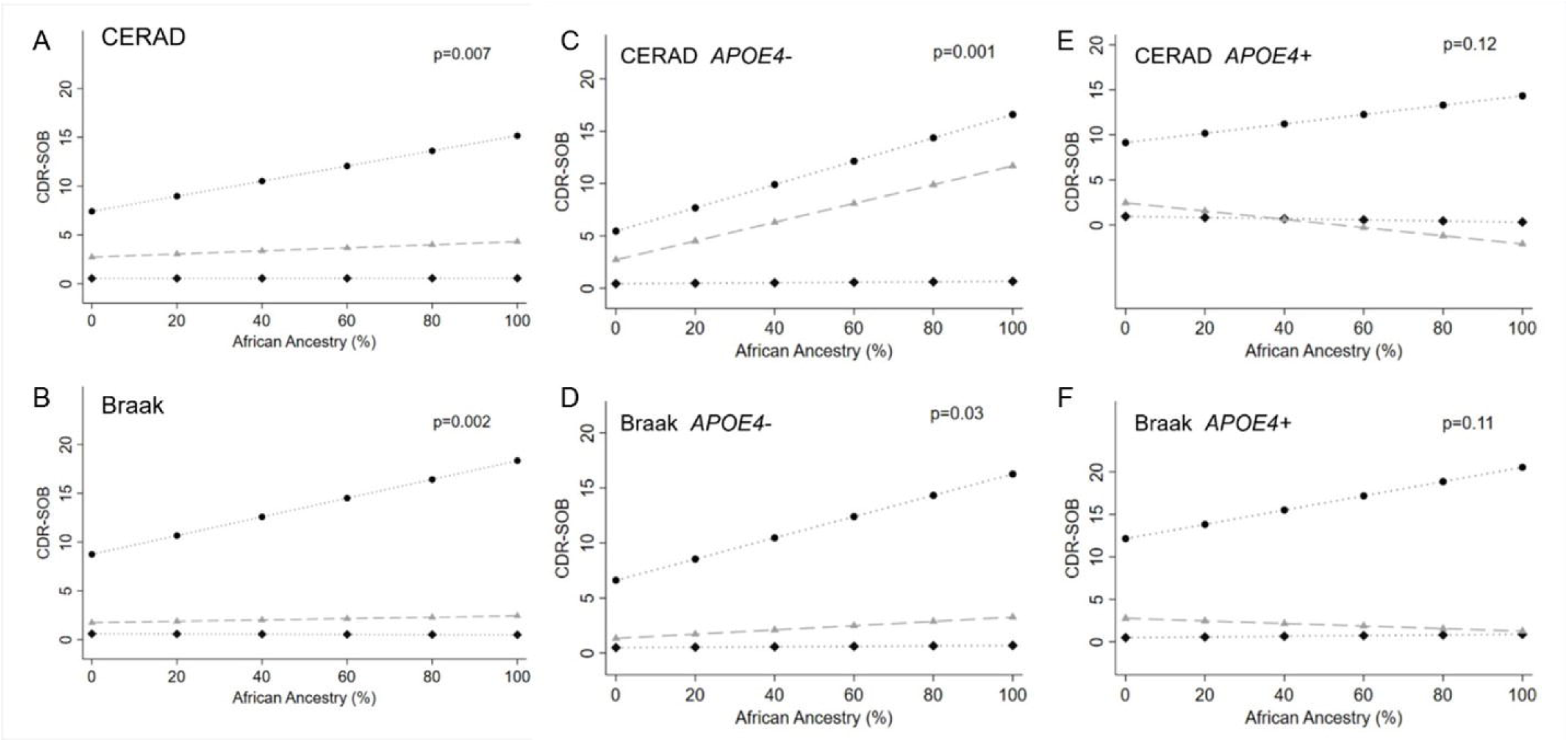
Association between African ancestry and Clinical Dementia Rating Sum of Boxes (CDR-SOB) considering an interaction term between African ancestry and neuritic plaques evaluated by the CERAD score (**A, C, and E**), or neurofibrillary tangle burden evaluated by the Braak & Braak stage (**B, D, and F**). (**A**) neuritic plaque burden in all individuals [Black diamond: None or sparse (n=303); Grey triangle: Moderate (n=49); Black circle: Frequent (n=48)]; (**B**) neurofibrillary tangle burden in all individuals [Black diamond: 0-II (n=263); Grey triangle: III-IV (n=99); Black circle: V-VI (n=38)]; (**C**) neuritic plaques in *APOE*4- individuals [Black diamond: None or sparse plaques (n=239); Grey triangle: Moderate plaques (n=31); Black circle: Frequent plaques (n=22)]; (**D**) neurofibrillary tangles in *APOE*4- individuals [Black diamond: Braak 0-II (n=206); Grey triangle: Braak III-IV (n=64); Black circle: Braak V-VI (n=22)]; (**E**) neuritic plaques in *APOE*4+ individuals [Black diamond: None or sparse plaques (n=64); Grey triangle: Moderate plaques (n=18); Black circle: Frequent plaques (n=26); and (**F**) neurofibrillary tangles in *APOE*4+ individuals [Black diamond: Braak 0-II (n=57); Grey triangle: Braak III-IV (n=35); Black circle: Braak V-VI (n=16)]. P-values for the interaction terms included in linear regression models adjusted for age, sex, education, and *APOE4* status (A and B). Neuritic plaques evaluated by the Consortium to Establish a Registry for Alzheimer’s disease (CERAD) score, and neurofibrillary tangles evaluated by the Braak staining system.

To further examine the impact of *APOE*4 in the association between cognitive abilities and AD-pathology according to AFR proportion, we tested and found a significant triple interaction between AFR plus *APOE*4 status with CERAD score (p=0.04), but not with Braak stages (p=0.77). Thus, we stratified individuals by *APOE*4 status to interrogate “Does the proportion of AFR impact the CDR-SOB scores independently of AD neuropathological burden as observed with the whole cohort without stratification?” Among *APOE*4- individuals, the results remained similar, meaning that the higher the AFR proportion, the worse the CDR-SB scores are in individuals with a higher burden of AD-neuropathology. However, the interactions lost significance in individuals *APOE*4+ (**Figure 2C-2F**).

### Local APOE4 ancestry

It is essential to recognize the importance of disconnecting global and local genomic ancestries regarding effect sizes in admixed populations. To further investigate a possible biological role of European vs. non-European *APOE* alleles into AD-neuropathology scores and cognitive abilities (CDR-SOB), we re-run regressions in a subgroup of 309 individuals stratified by their *APOE* status (*APOE*4+ and *APOE*4-) and local ancestry (EUR vs. non-EUR), either adjusting or not for global ancestry (**Figure 1**).

First, we tested the *APOE*4*-*group (**Table 3**) because we only observed worse CDR-SOB scores associated with a higher global AFR proportion in this group (**Figure 2C and 2D**). Compared to individuals with local EUR *APOE*4*-* genotypes (homozygous for local ancestry), those with local non-EUR *APOE*4*-*genotypes had lower CERAD scores independent of the adjustment for global AFR (**Table 3**), in line with results obtained when considering global ancestry only. Despite the lower odds to accumulate neuritic plaques, the CDR-SOB scores were worse in the local non-EUR *APOE*4*-*groups when adjusting only for the CERAD scores, meaning that once plaques accumulate, the functional outcome seems to be worse in non-EUR *APOE*4*-* individuals (mixed plus non-EUR *APOE*4*-* alleles, see **Figure 1**), exactly as observed in the global ancestry only analyses. This association lost statistical significance after correcting for global AFR proportion, although the trend remains (p=0.059).

**Table 3.** Association between AD neuropathological burden and functional cognitive scores with one or two non-European *APOE* alleles (local APOE ancestry) in *APOE4-* individuals (n=235, see Figure 1 for grouping criteria)

| Outcomes | Pathology adjustments | Model 1 (without AFR adjustment) |  | Model 2 (with AFR adjustment) |  |
| --- | --- | --- | --- | --- | --- |
| | | $\beta$ (95% CI) | p | $\beta$ (95% CI) | p |
| **BB** | NA | -0.12 (-0.46; 0.22) | 0.493 | -0.23 (-0.63; 0.17) | 0.263 |
| **CERAD** | NA | -0.27 (-0.48; -0.05) | **0.015** | -0.28 (-0.53; -0.02) | **0.032** |
| **CDR-SOB** | NA | 0.35 (-0.72; 1.42) | 0.518 | 0.35 (-0.92; 1.62) | 0.585 |
|  | BB+CERAD | 0.89 (-0.07; 1.86) | 0.069 | 1.00 (-0.14; 2.15) | 0.084 |
|  | BB | 0.46 (-0.52; 1.43) | 0.358 | 0.53 (-0.63; 1.69) | 0.372 |
|  | CERAD | 1.01 (0.04; 1.99) | **0.041** | 1.11 (-0.04; 2.26) | 0.059 |

Reference: Individuals EUR *APOE4*- alleles (genotypes 22, 23 or 33) (n=138)
AFR: African ancestry (continuous 10% increments); BB: Braak & Braak; CERAD: Consortium to Establish a Registry for Alzheimer's disease; CDR-SOB: Clinical Dementia Rating sum of boxes, NA: not applicable
Model 1: Linear regression model adjusted for age, sex, education, and AD neuropathology when indicated.
Model 2: Linear regression model adjusted for age, sex, education, AD neuropathology when indicated, and AFR.

As we hypothesize that the *APOE4* allele has an attenuated effect in non-EUR ancestry, we compared the effect magnitude in *APOE4-* vs. *APOE4+* in individuals with local EUR in *APOE* separately from individuals with non-EUR *APOE* (see **Figure 1** groups). We excluded individuals with a mix of EUR and non-EUR *APOE* alleles for this analysis. Table 4A shows the results for individuals with EUR *APOE* (**Table 4A**, *APOE4+* vs. *APOE4-*, **Figure 1 groups A1 vs. B1**). As expected, EUR *APOE4+* individuals had higher Braak stages than EUR *APOE4-*, even after adjusting for global ancestry; interestingly, the CDR-SOB scores were similar for both groups, even after adjusting for the burden of AD-pathology.

**Table 4.** Association of AD neuropathological burden and functional cognitive scores with local European or non-European *APOE*4+ ancestries using corresponding ancestries of *APOE*4- individuals as references.

| **(A) EUR *APOE4*+ local ancestry (n=31)\*** |  | **Model 1 (no AFR adjustment)** |  | **Model 2 (with AFR adjustment)** |  |
| --- | --- | --- | --- | --- | --- |
| **Outcomes** | **Pathology adjustments** | **β (95% CI)** | **p** | **β (95% CI)** | **p** |
| **BB** | NA | 0.52 (0.01; 1.03) | **0.048** | 0.51 (-0.002; 1.03) | 0.051 |
| **CERAD** | NA | 0.61 (0.28; 0.95) | **<0.001** | 0.61 (0.27; 0.95) | **<0.001** |
| **CDR-SOB** | NA | 1.22 (-0.49; 2.93) | 0.16 | 1.29 (-0.41; 2.99) | 0.14 |
|  | BB+CERAD | 0.52 (-1.06; 2.10) | 0.52 | 0.55 (-1.02; 2.13) | 0.49 |
|  | BB | 0.92 (-0.66; 2.50) | 0.25 | 0.97 (-0.61; 2.54) | 0.23 |
|  | CERAD | 0.25 (-1.34; 1.83) | 0.76 | 0.31 (-1.27; 1.89) | 0.70 |
| **(B) Non-EUR *APOE4*+ local ancestry (n=17)\*\*** |  | **Model 1 (no AFR adjustment)** |  | **Model 2 (with AFR adjustment)** |  |
| **BB** | NA | -0.21 (-1.08; 0.65) | 0.62 | -0.21 (-1.10; 0.67) | 0.62 |
| **CERAD** | NA | 0.01 (-0.52; 0.70) | 0.76 | 0.09 (-0.51; 0.69) | 0.76 |
| **CDR-SOB** | NA | 0.39 (-2.78; 3.56) | 0.80 | 0.39 (-2.82; 3.60) | 0.81 |
|  | BB+CERAD | 1.53 (-1.60; 4.66) | 0.33 | 1.53 (-1.66; 4.72) | 0.33 |
|  | BB | 1.45 (-1.41; 4.32) | 0.31 | 1.45 (-1.46; 4.37) | 0.32 |
|  | CERAD | 0.59 (-2.50; 3.67) | 0.70 | 0.59 (-2.54; 3.73) | 0.70 |

\*Reference for (A): Individuals *APOE4*- (genotypes 22, 23, or 33) with local European ancestries (n=138)
\*\* Reference for (B): Individuals *APOE4*- (genotypes 22, 23 or 33) with local non-European ancestries (n=24)
AFR: African ancestry (continuous 10% increments); BB: Braak & Braak; CERAD: Consortium to Establish a Registry for Alzheimer's disease; NA: not applicable
Model 1: Linear regression model adjusted for age, sex, education, and AD neuropathology when indicated.
Model 2: Linear regression model adjusted for age, sex, education, AD neuropathology when indicated, and AFR.
OBS. Only individuals homozygous for *APOE* local ancestry were included

Next, we tested the local non-EUR *APOE*4+ individuals (**Table 4B**, Non-European *APOE4+* vs. *APOE4-*, **Figure 1 groups A4 vs. B3**). As opposed to individuals EUR at the *APOE* locus, we did not find associations of local ancestry with AD-pathology (nor with cognitive scores). Results did not change after adjustment for global ancestry. Results from Table 4 support the hypothesis that the effect of *APOE4+* into neuropathological AD burden is attenuated in non-EURs when using corresponding local ancestry *APOE4-* individuals as references.

## Discussion

Dementia differentially affects individuals based on race due to a combined contribution of ancestry-based genetic factors and social determinants of health ^1, 2^. Studies with comprehensive neuropathological and DNA-based ancestry assessments in large cohorts with admixed individuals have the potential to help disentangle the nature vs. nurture contributions to these differences. Here, we analyzed 400 individuals with a broad range of AD neuropathological severity and absence of other significant neuropathological changes from a clinical-neuropathological-genetic cohort of 1,333 individuals. Ancestry proportions measured by DNA analysis unveiled a high admixture of EUR and AFR ancestry with a modest Brazilian Native American ancestry component. Validating and expanding from our previous study ^22^, we show that AFR correlates with a lower burden of neuritic plaques even after adjusting for several factors, including *APOE* status. Next, as we found an interaction among AFR, CERAD scores for neuritic plaques, and *APOE* genotypes, we interrogated if *APOE*4 conferred a similar risk to individuals of AFR compared to EUR ancestry.

We found that among *APOE*4*-* individuals with a severe burden of neuritic plaques and tau neurofibrillary tangles, the higher the AFR proportion, the worse the functional cognitive scores. This observation mainly persisted upon stratification by local *APOE* ancestry after adjusting for neuritic plaques burden, independent of global ancestry status, even though the CERAD outcome continued to present the opposite effect (less severity in AFR). Paradoxically, we found no association between the proportion of AFR and AD metrics in the *APOE*4+ group. Further analysis comparing the magnitude effect of *APOE*4*+ vs. APOE*4*-* in neuropathological and functional outcomes in local *EUR vs. AFR* APOE groups showed a negative impact of *APOE*4*+ (*worse AD neuropathological metrics than in *APOE*4*-* individuals) in EUR only. These combined results corroborate the hypothesis of an attenuated effect of non-EUR *APOE*4*+* in AD neuropathology.

The literature shows a higher prevalence of dementia in Blacks than in Whites ^33^, but most of these studies analyzed cohorts lacking neuropathological assessment and/or DNA-based ancestry determination. Thus, non-AD neuropathological changes, such as cerebrovascular changes, a known contributor to cognitive decline and more prevalent in non-EUR, make it difficult to interrogate the relationship of risk factors to AD neuropathological changes. Our study suggests that the burden of AD neuropathology is not likely to explain this difference in cognitive scores. Moreover, since our cohort lacks noticeable non-AD neuropathological changes, it is also unlikely that comorbid neuropathological changes explain the differences in cognitive outcomes. Despite our best efforts to eliminate other confounders: using a cohort of individuals dwelling in the same city and adjusting the models for education, a proxy of social determinants of health, we cannot exclude that a combination of social factors contributed to a worse cognitive reserve in individuals with increasing AFR proportion, which is contributing for the worse cognitive scores ^34-36^. A study from Cuba shows worse cognitive scores in individuals with a high AFR proportion, only before adjusting for socioeconomic factors ^37^. Although this study is not directly comparable to ours (no neuropathology assessment and dichotomous classification of AFR), it shows how social determinants of health enriched in specific populations may impact cognition. In addition to sociodemographic factors, *APOE*2 (ε2 allele) and *APOE*3 (ε3 allele) effects on cognition may differ across ancestries. *APOE*2 is generally considered to have a protective effect against AD in EUR ancestry, but less is known in non-EUR ancestry. Weaker APOE2 effect size was reported recently in non-EUR local ancestry context ^8^. In a multi-racial cohort of New York City, *APOE*2/3 genotype was associated with an 8x increased risk of dementia of AD-type in African-Americans (self-declared race). However, it was associated with reduced risk in Whites ^38^. Unfortunately, *APOE* genotypes ε2/ε2 and ε2/ε3 are less common genotypes, which challenges robust association studies due to sample size limitations, particularly in detailed neuropathology-based studies in population-representative samples such as ours. Further studies should interrogate if a poor protective effect of *APOE*2 in non-EUR contributes to the worse functional cognitive scores in this group.

The most intriguing observation of this study is the attenuation of the relationship between higher AFR proportion and worse CDR-SOB scores among *APOE4* carriers and the attenuation of AD neuropathology burden in *APOE*4 carriers vs. noncarriers in individuals with non-EUR *APOE* background. Including local ancestry analysis is critical in admixed population studies because global ancestry is inferred, grouping a small number of ancestry-informative markers. Thus the local ancestry of a given gene may be different from the global ancestry estimates (i.e EUR/EUR or EUR/non-EUR local ancestry in an individual with a predominant non-EUR global estimate and vice-versa).

The literature also suggests a possible biological attenuation of risk AD-related cognitive decline among *APOE*4 carriers of AFR vs. EUR ancestry. In a comparison performed by Rajabli and colleagues ^6^, of AD cases and controls (no neuropathological confirmation for any group) of African American and Puerto Rican populations, *APOE* presented a differential effect based on AFR in admixed individuals. Although *APOE*4 is a significant risk allele for AD in both populations, odds ratio was 1.3-3.5-fold higher in EUR backgrounds ^6^. Blue and colleagues showed that *APOE*2 and *APOE*4 had weaker effects (although in the same direction) in Caribbean Hispanics clinical AD (dementia of AD-type, no neuropathological assessment) cohorts compared with a cohorts of EUR with dementia of AD-type ^8^. Notably, *APOE*4 homozygosity was the main contributor to AD hazard ratio, as expected, but AFR local *APOE* decreased the hazard ratio by 28% ^8^. Another study suggests a similar effect of *APOE*4 in AFR and EUR (global ancestry), but unfortunately, this study lacked neuropathological assessments ^11^.

Interestingly in the same study, the effect of *APOE*4 is attenuated in the subgroup of Dominicans, the closest in ancestry composition as compared to our Brazilian cohort. Furthermore, a study from New York found that a higher risk of dementia among African Americans and Hispanics was not related to *APOE4* ^39^. Individuals with the *APOE*4 allele get an advantage in high-pathogen and energy-limited contexts by experiencing reduced innate inflammation and maintaining higher lipid levels ^36, 40^. The proportion of *APOE*4 in cognitively normal AFR individuals is higher than in other populations^41^. Complex interactions between cognitive outcomes and cardiovascular phenotypes such as triglyceride levels ^42^, arterial thromboembolism, aneurism, and obesity ^43^ may contribute to attenuated *APOE*4 effect in individuals with AFR ancestral ancestry.

*APOE*4 mRNA expression levels are higher in EUR than in AFR allele ^44^, which could explain the larger effect-size. Interestingly, a recent single-cell expression study suggested that frontal cortex cells of AD patients significantly overexpress *APOE*4 in EUR local APOE ancestries than in AFR local ancestries ^44^. This observation corroborates with the previous findings and offers a local ancestry-specific *APOE* expression regulation hypothesis and favoring the existence of a polygenic modulation of *APOE* net penetrance that can be proxied by global ancestry in admixed individuals. Moreover, haplotypic structure of population-specific rare and common variant combinations may underlie the attenuation of *APOE*4 deleterious effect in AFR. Indeed, relevant functional haplotypes may include neighboring genes such as *TOMM40*, also associated with AD ^45^, pointing that ancestry-specific effects from neighboring genes may be driving our findings ^6^. Local ancestry was also significantly associated with known AD-related loci such as *ABCA7* and *CD33*, and loci without previous association with AD such as *CYP4B1, DAB1, MYSM1* and others, always controlling for *APOE* genotypes ^46^.

This study has several strengths, including a community-dwelling sample from an admixed population, neuropathological assessment, exclusion of individuals with non-AD neuropathological changes, DNA-based ancestry determination, and known *APOE* background. However, it is important to point out its limitations. Despite having one of the largest cliniconeuropathological series with admixed individuals, the number of individuals with *APOE*4+ genotypes is small, especially with the non-EUR background. Also, we run multiple tests. In combination, it increases the chances of error type I or II, although our results align with the literature of clinical samples. Also, we used semi-quantitative rather than quantitative neuropathological assessment and two different methods to input *APOE* genotype as the collection spans several years. However, quality control comparisons showed a similarity of results with any of the methods.

In conclusion, our work corroborates the need to increase the number of admixed populations in AD research. It suggests that neuropathology, clinical outcomes, and APOE genotyping differ with different ancestry backgrounds. The association between between increased AFR proportion and worse functional cognitive scores was lost in *APOE*4+, supporting the hypothesis that *APOE*4 risk in AD is attenuated in individuals with AFR ancestry compared to EUR ancestry.

## Data Availability

All data produced in the present study are available upon reasonable request to the authors

## Acknowledgments

We are grateful to the families who donated brains to the Biobank of Aging Studies of the University of Sao Paulo and the Sao Paulo Autopsy Service for the partnership. This work received funding from Sao Paulo Research Foundation (grant numbers 06/55318-1, 09/09134-4, 13/08028-1, 14/50931-3 16/24326-0; 18/16626-0) Conselho Nacional de Desenvolvimento Científico e Tecnológico (CNPq) (466763/2014-0, 465355/2014-5, 403502/2020-9), Alzheimer Association and National Institute of Health (K24AG053435), FAPEMIG (00314-16). Part of the genetic data was generated with funds from NIH R01AG17917.

