## Supplementary Information for "Global and local ancestry modulate *APOE* association with Alzheimer’s neuropathology and cognitive outcomes in an admixed sample"

### Supplementary methods

#### DNA extraction

DNA samples were extracted from post-mortem cardiac blood samples using Gentra Autopure LS automated protocol, followed by measurement of DNA quality and concentration spectrophotometer, as previously described [1]. If DNA parameters have not reached minimum standards for genetic analyses, DNA was isolated and purified from frozen cerebellum using manual standard salt and EDTA cell lysis followed by phenol-chloroform isoamyl alcohol extraction yielding adequate samples for subsequent analyses.

#### Global ancestry inference

BAS samples used in this study were subjected to either one or both of the following genotyping methods. A total of 724 individuals were genotyped using Illumina Human OmniExpress 700k microarray, of which 309 were included in this study. The remaining 91 individuals were genotyped using the Illumina BeadXpress platform, using a custom panel containing ancestry informative markers (AIMs), used to calculate global ancestry [2]. To maximize the number of samples and avoid the imputation of the non-overlapping variants, global ancestry inference was performed with genotypes of the overlapping 47 SNVs. The global ancestry correlation of this subset of markers and the full panel was 0.94, after verification using 246 samples that were genotyped using both the microarray and the custom BeadXpress panel.

We inferred sample structure with Structure 2.3.4 [3]. The run was performed assuming K=3 populations, based on the main parental population groups that originated modern Brazilians [4]; additional run parameters included a burn-in procedure of 100,000 steps, followed by 100,000 Markov Chain Monte Carlo iterations, using the admixture model with allele frequency correlated among populations. In order to assist ancestry inference, we included genotypes of individuals from reference populations, obtained from the HapMap Project (Central Europeans from Utah, n=57; Yoruba in Ibadan, Nigeria, n=62) and Kosoy et al. (2009) [2]; 188 European descent from Utah and New York, 98 West African individuals from Nigeria and Niger-Congo region and 105 Native American descent individuals from Guatemala, Peru, and Mexico).

#### *APOE* genotyping, imputation, and local ancestry inference

*APOE* common alleles are composed of a combination of genotypes in variants rs7412 and rs429358, the latter of which is absent from the microarray. Imputation was performed using a window from the genotyping array dataset composed of 724 individuals (including the 309 used in this study) to determine the rs429358 genotype. The window included 2.216 SNPs in a ~10Mb (11.323.329 bp) region from 40.404.633 bp to 51.727.962 bp from chromosome 19, which includes the rs7412 (19:45412079) but not the rs429358 (19:45411941) in GRCh37 assembly. This region has been selected to focus on ~±5 Mb around the rs7412 interest.

We imputed the target dataset with a reference panel that merged the public reference panel data from 1000 Genomes Phase 3 Project (1KGP) and 270 individuals from EPIGEN-Brazil (90 of each cohort) genotyped for 4.3 million markers, the EPIGEN-5M+1KGP imputation panel, fully described in Magalhães et al. (2018) [5] and considered only SNPs imputed with an info score quality metric > 0.8. All imputation tasks detailed below were performed using the master script described elsewhere [5]. Pre-phasing between the target and reference panels has been done using SHAPEIT2 [6] to check the consistency of the marker's strand of target and reference panels with the human genome reference sequence and PLINK software [7] to flip the strands in case of inconsistencies. Then, haplotype phase inference of the target dataset has been made using the 1KGP haplotypes. Finally, imputation has been done using the EPIGEN-5M+1KGP imputation panel, and IMPUTE2 v.2.3.2 software [8] on chromosome chunks of 7 Mb, with effective size parameter (Ne) set to 20,000 and the IMPUTE2 info score as a metric of imputation quality. QCTOOL (Available at: https://www.well.ox.ac.uk/~gav/qctool/) has been used for data quality control, filtering, conversions.

Imputation performance was verified with direct *APOE* genotyping obtained using allele-specific amplification and real-time PCR, as described previously [9], which was conducted orthogonally in BAS samples, including most that were genotyped using microarray and custom panel. For a total of 523 cases that overlapped *APOE* genotyping methods, we obtained a 93.3% accuracy and chose the result from direct genotyping in all samples available. Among the 309 individuals with genotyped array included in this study, a total of 28 cases were APOE genotyped using imputation of rs429358.

For local ancestry inference (LAI), we selected Africans, Europeans, Native Americans from Public datasets (1000Genomes Project [10] and Human Genome Diversity Project [HGDP] [11]

WGS). Specifically, from 1000 Genomes, we selected YRI and LWK as African references, CEU and IBS as European. For the Native American ancestry, we selected individuals with a high proportion of Native American Ancestry from PEL and MXL and Native Americans from HGDP, in addition to other Native American samples recently published [12].

We selected the genomic region corresponding to the *APOE* gene (GRCh37, chromosome 19 from 40416921 to 51727962, ~11Mb), including 2,234 SNPs in our dataset. We merged with the public datasets using PLINK, generating a dataset with 1349 individuals. We phased the merged data using SHAPEIT2[6]. We applied RFMix [13] using two Expectation-Maximization iterations with a minimum node size of 5. Only inferred local ancestries per haplotype with 80% or above posterior probabilities were included.

**Supplementary Figure**

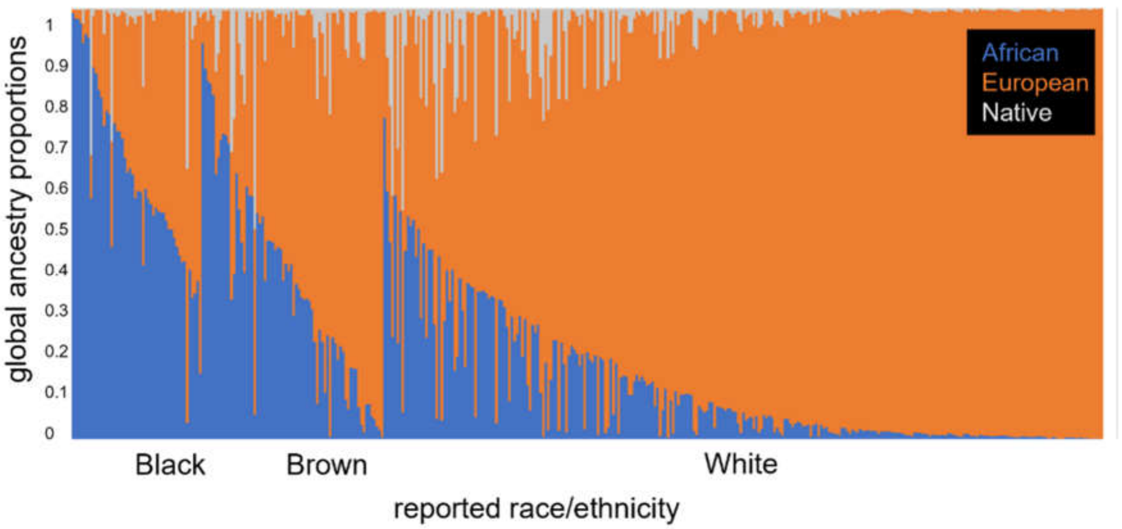


**Supplementary Figure 1.** Distribution of global ancestry proportions vs. reported race/ethnicity in the study population (N=397, three individuals of Asian reported race/ethnicity not shown in graph).

**Supplementary Table 1**. Distribution of ancestry proportions across reported race/ethnicity groups

| | | Ancestry proportions (average ± stdev [min-max]) | | |
|---|---|---|---|---|
| **Reported race/ethnicity** | **N*** | **EUR** | **AFR** | **NAT** |
| **Black** | 50 | 0.36 ± 0.2 [0.01-0.83] | 0.61 ± 0.22 [0.05-0.99] | 0.04 ± 0.09 [0.01-0.38] |
| **Brown** | 70 | 0.59 ± 0.24 [0.07-0.99] | 0.36 ± 0.25 [0.01-0.92] | 0.06 ± 0.09 [0.01-0.51] |
| **White** | 277 | 0.86 ± 0.17 [0.25-1] | 0.11 ± 0.14 [0.01-0.75] | 0.04 ± 0.08 [0.01-0.47] |
| **All** | 397 | 0.75 ± 0.26 [0.01-1] | 0.22 ± 0.25 [0.01-0.99] | 0.05 ± 0.09 [0.01-0.51] |

*Three samples reported as Asians were excluded from the ancestry vs. reported race/ethnicity distribution description

**Supplementary Table 2. Association between dichotomous African ancestry* and AD neuropathological burden (n=400)**

| | Crude | | Model 1 | | Model 2 | |
|---|---|---|---|---|---|---|
| | **β (95% CI)** | **p** | **β (95% CI)** | **p** | **β (95% CI)** | **p** |
| **BB stage** | 0.077 (-0.308; 0.461) | 0.70 | 0.147 (-0.172; 0.465) | 0.37 | 0.137 (-0.177; 0.450) | 0.39 |
| **CERAD score** | -0.233 (-0.477; 0.010) | **0.06** | -0.197 (-0.416; 0.021) | **0.08** | -0.210 (-0.418; -0.001) | **0.04** |

*African ancestry was considered positive if equal or greater than 2% of African ancestry.
BB: Braak & Braak; CERAD: Consortium to Establish a Registry for Alzheimer's disease.
Model 1: Linear regression model adjusted for age, sex, and education.
Model 2: Linear regression model adjusted for age, sex, education, and *APOE4* status.
